# Vaccine effectiveness with BNT162b2 (Comirnaty, Pfizer-BioNTech) vaccine against reported SARS-CoV-2 Delta and Omicron infection among adolescents, Norway, August 2021 to January 2022

**DOI:** 10.1101/2022.03.24.22272854

**Authors:** Lamprini Veneti, Jacob Dag Berild, Sara Viksmoen Watle, Jostein Starrfelt, Margrethe Greve-Isdahl, Petter Langlete, Håkon Bøås, Karoline Bragstad, Olav Hungnes, Hinta Meijerink

## Abstract

**Background:** COVID-19 vaccination was recommended for adolescents in Norway since August 2021. In this population-based cohort study, we estimated the BNT162b2 vaccine effectiveness against any PCR-confirmed (symptomatic or not) SARS-CoV-2 infections caused by the Delta and Omicron variant among adolescents (12-17-years-old) in Norway from August 2021 to January 2022.

**Methods:** Using Cox proportional hazard models, we estimated the BNT162b2 vaccine effectiveness against any Delta and Omicron infections. Vaccine status was included as a time-varying covariate and models were adjusted for age, sex, comorbidities, county of residence, country of birth, and living conditions. Data were obtained from the National Preparedness registry for COVID-19, which contains individual-level data from national health and administrative registries.

**Findings:** Vaccine effectiveness against Delta infection peaked at 68% (95%CI: 64–71%) and 62% (95%CI: 57– 66%) in days 21-48 after the first dose among 12–15-year-olds and 16–17-year-olds respectively. Among 16–17-year-olds that received two doses, vaccine effectiveness peaked at 93% (95%CI: 90–95%) in days 35-62 and declined to 84% (95%CI: 76–89%) in 63 days or more after the second dose. For both age-groups, we found no protection against Omicron infection after receiving one dose. Among 16–17-year-olds, vaccine effectiveness against Omicron infection peaked at 53% (95%CI: 43–62%) in 7-34 days after the second dose and decreased to 23% (95%CI: 3–40%) in 63 days or more after vaccination. Vaccine effectiveness decreased with time since vaccination for both variants, but waning was observed to occur faster for Omicron.

**Interpretation:** Our results suggest reduced protection from BNT162b2 vaccination against any SARS-CoV-2 infection caused by the Omicron variant compared to the Delta. In addition, waning immunity was observed to occur faster for Omicron. The impact of vaccination among adolescents on reducing infection and thus transmission is limited during omicron dominance.

**Funding:** No funding was received.

**Research in context:** 

**Evidence before this study:** BNT162b2 (Comirnaty, Pfizer-BioNTech) and mRNA-1273 (Spikevax, Moderna) vaccines have been approved for use in adolescents, based on results from randomized placebo-controlled trials demonstrating comparable immunogenicity and safety profile as in young adults. In addition, observational studies from Israel, the USA and England have reported high protection of BNT162b2 vaccines against SARS-CoV-2 Delta infection among adolescents. These studies also reported decrease in effectiveness with time since last vaccine dose. Evidence on the effect of an extended interval between doses, longer time since vaccination and the effect against different variants is limited. When we first planned this study in early February 2022, no data were available regarding vaccine effectiveness against SARS-CoV-2 Omicron infection among adolescents. To our knowledge when we completed this study and before submitting this article, only one study from England reported results in a preprint on vaccine effectiveness against symptomatic SARS-CoV-2 Omicron infection among adolescents. We searched for studies that evaluated vaccine efficacy or effectiveness after vaccination of adolescents during 2021-2022 in PubMed, medRxiv, bioRxiv, SSRN. We searched for studies with several variations of the primary key search terms “COVID-19”, “SARS-CoV-2”, and “vaccine” (including names of specific vaccines, as BNT162b2), “vaccine effectiveness”, “adolescents”, “children”.

**Added value of this study:** The rapid increase in the incidence of SARS-CoV-2 infection caused by the Omicron variant in highly vaccinated populations has raised concerns about the effectiveness of current vaccines in adults but also adolescents. In this population-based cohort study, we showed that the vaccine effectiveness against Omicron is lower than against Delta infections among adolescents, including symptomatic and asymptomatic infections. We should note that evidence suggests higher rates of asymptomatic carriage for Omicron than other variants of concern. Vaccine effectiveness that includes asymptomatic cases, as in the study from England, is expected to be lower than when including symptomatic cases only. We found that one and two doses of BNT162b2 among adolescents protected well against Delta. Vaccination provided high protection against Delta infections (>91%) among Norwegian 16-17-year-olds 7-62 days after the second dose. We found no protection against Omicron SARS-CoV-2 infection after one vaccine dose, and moderate effectiveness after two doses (peaked at 53%) among the 16-17-year-olds. Moreover, waning immunity was observed to occur faster for Omicron.

**Implications of all the available evidence:** Based on the available evidence, the vaccine effectiveness among adolescents is similar to that reported among adults, also with an extended period of 8-12 weeks between doses which was used in Norway. Protection is significantly lower against Omicron than Delta infections and immunity wanes faster against Omicron. The impact of vaccination among adolescents on reducing infection and thus transmission is limited during omicron dominance. Policies should take into account the impact of vaccination campaigns among adolescents and their primary objective. Vaccine effectiveness should be re-evaluated when other variants appear as they might have different outcomes as shown between Delta and Omicron infections.

## Background

Even though COVID-19 is generally mild in children of all ages (1, 2), strict and widespread infection prevention measures to control transmission in society have disrupted their education and everyday social activities (3).To provide advice for infection control among children and adolescents, including vaccination, it is essential to understand the role of children in transmission, their disease burden as well as the effect and consequences of implemented measures.

Overall, vaccines are considered an important measure to fight the spread the COVID-19 and prevent serious outcomes. Since the end of 2020, several COVID-vaccines have been granted conditional marketing authorization for use in adults by the European Medical Agency (EMA). In addition, BNT162b2 (Comirnaty, Pfizer-BioNTech) and mRNA-1273 (Spikevax, Moderna) vaccines have been approved for use in adolescents, based on results from randomized placebo-controlled trials demonstrating comparable immunogenicity and safety profile as in young adults (4, 5). The effectiveness of BNT162b2 in adolescents has also been demonstrated in observational studies from Israel, the USA, and England, showing very good protection against infection with the Delta variant after the second dose (6-9). The effectiveness of one dose and of the second dose among adolescents against symptomatic Omicron infection has been explored in a study from England published as a preprint, but the evidence is still limited (9).

Since 18 August 2021, Norway has recommended two doses of BNT162b2 for adolescents aged 16-17 years, with an extended interval of 8-12 weeks between doses. Additionally, since 2 September 2021, adolescents between 12 and 15 years were offered a single dose of BNT162b2, and a second dose from the end of January 2022 (10). From mid-August (at school re-entry), testing in secondary schools was widespread and lasting throughout the study period. All class contacts of cases were encouraged to test, and in September additionally routine biweekly testing in secondary schools was introduced in areas and periods of high incidence. All testing was free of charge and easily accessible to ensure a high uptake. Most adolescents 12-17 years would be tested using rapid antigen test, followed by a PCR test to confirm if positive. In Norway, the Delta variant was predominant from August to December 2021. The Omicron variant was first detected in adults at the end of November and overtook Delta by the end of the year (11, 12). The rapid increase in the incidence of SARS-CoV-2 infection caused by the Omicron variant in highly vaccinated populations has raised concerns about the effectiveness of current vaccines in adults but also adolescents.

In this study, we estimated the vaccine effectiveness of BNT162b2 against any PCR-confirmed (symptomatic or not) SARS-CoV-2 Delta or Omicron infection, considering time since last dose among adolescents between the ages of 12-17 years in a population-based cohort study in Norway from August 2021 to January 2022. The results from this study will allow us to evaluate the duration of protection among adolescents vaccinated with BNT162b2, which is necessary to guide future vaccination policy.

## Methods

### Study design and data sources

We conducted a retrospective population-based cohort study for the period 25 August 2021 to 16 January 2022 including all individuals born from 2004 to 2009 (12-17 year old), who were registered as living in Norway with a valid national identity number. We excluded 4,374 individuals for which the interval between first and second dose was shorter than the minimum interval (19 days), those with four registered vaccine doses, or who received another vaccine product (not BNT162b2). In addition, 14,439 individuals with a SARS-CoV-2 infection reported prior to 25 August 2021 were excluded.

We obtained data from the Norwegian national preparedness registry for COVID-19 (13). The preparedness registry contains individual-level data from central health registries, national clinical registries, and other national administrative registries, which can be linked using the national identification number. It covers all residents in Norway and includes data on all laboratory-confirmed cases of COVID-19, all hospitalisations among cases and COVID-19 vaccinations (supplementary materials, part 1). Individual-level data used for this study included age (in years), sex, county of residence (12 levels), household crowding (three levels; yes, no, unknown), dates of vaccination, vaccine type (only BNT162b2 included), underlying comorbidities (three levels), date of sampling for reported cases and information on SARS-CoV-2 variant if available (supplementary materials, part 1). We extracted data from the registries on 18 February 2022.

### Definitions

#### SARS-CoV-2 infection

We defined SARS-CoV-2 infection as a positive SARS-CoV-2 PCR test reported to the Norwegian Surveillance System for Communicable Diseases (MSIS) registry. We use testing date as time of infection (positive PCR test). Both symptomatic and asymptomatic reported cases have been included as it is not possible to distinguish between these in MSIS.

#### Vaccination status

Vaccine status was defined based on number of doses and date of vaccination recorded in the Norwegian Immunisation Registry (SYSVAK) as:

1. Unvaccinated: no dose received
2. One dose, 0-20 days after first vaccine dose
3. One dose, 21-48 days after first vaccine dose
4. One dose, 49-76 days after first vaccine dose
5. One dose, ≥77 days after first vaccine dose
6. Two doses, 0-6 days after second vaccine dose
7. Two doses, 7-34 days after second vaccine dose
8. Two doses, 35-62 days after second vaccine dose
9. Two doses, ≥63 days after second vaccine dose

#### Underlying comorbidities with increased risk of severe COVID-19

Individuals with underlying comorbidities that cause increased risk of severe COVID-19 have been prioritized for vaccination. We categorised individuals into three groups (details in supplement part 1): i) low risk (no underlying comorbidities), ii) medium risk comorbidity and iii) high risk comorbidity.

### Laboratory methods

SARS-CoV-2 variants were identified based on whole genome sequencing, Sanger partial S-gene sequencing, or PCR screening targeting specific single nucleotide polymorphisms, insertions or deletions that reliably differentiate between Omicron and other variants. The laboratory testing for variants of SARS-CoV-2 in Norway has been described in further detail elsewhere (12). We included data on reported cases of laboratory-confirmed SARS-CoV-2 infection from MSIS. As of January 2022, reinfections are registered in MSIS if there are ≥6 months between two positive sampling dates for an individual.

### Data analysis

#### Statistical analysis

We estimated the vaccine effectiveness against any SARS-CoV-2 infections (Delta- and Omicron infections in separate analyses), using a Cox proportional hazards models with vaccination status as a time-dependent covariate, and with explicit time to account for changes in the baseline hazard over time. In these analyses we stratified for available factors associated with the likelihood of being vaccinated and being infected. In all our adjusted vaccine effectiveness estimates presented here, we stratified for sex, country of birth, county of residence, crowding and underlying comorbidities associated with increased risk of severe COVID-19. Vaccine effectiveness is defined as 100*(1 –β), with β the proportional hazard associated with vaccine status.

More specifically, we estimated the vaccine effectiveness among individuals with no prior SARS-CoV-2 infection after receiving one dose for individuals 12-15 years old and after one or two doses for individuals 16-17 years old against a) all reported infections from 25 August to 25 November, b) reported Delta infections from 25 August 2021 to 16 January 2022, and c) reported Omicron infections from 26 November to 16 January. The analyses for all reported cases during the period 25 August to 25 November is conducted assuming that all reported cases were Delta since Delta accounted for nearly 100% of all samples analysed for genetic variant in that period. We restricted the vaccine effectiveness analyses for Omicron infection after the first case of Omicron was reported in adults on the 26 of November 2021. We closed the follow up period on 16 January since the comprehensive screening activity in Norway ceased as Omicron reached more than 90% prevalence.

In addition, in supplement part 2 we present sensitivity analyses that were performed to evaluate the impact in estimating vaccine effectiveness for a specific variant when only including (using data) on screened cases since usually only part of all reported cases is screened for variant.

Statistical analysis was performed in Stata version 16 (Stata Corporation, College Station, Texas, US).

#### Ethics approval

Ethical approval was granted by Regional Committees for Medical and Health Research Ethics (REC) Southeast (reference number 122745).

#### Role of the funding source

This research did not receive any specific grant from funding agencies in the public, commercial, or not‒for‒ profit sectors. The study was performed as part of routine work at the Norwegian Institute of Public Health.

## Results

Overall, 372,179 adolescents 12-17 years old were included in this cohort study and 52,891 of those were reported to have been infected with COVID-19 cases (first infections) by the end of the study period. Out of the 52,891 reported cases, 42% were screened for variants and from those 26% of variants were sequenced. The first Omicron case in adolescents 12-17 years old was detected on the 2^nd^ of December 2021 (week 48). The proportion of cases screened for variants varied throughout the study period, with the highest proportion (67%) in week 50 following the period after the detection of the first Omicron cases. Variant detections ranged from 25 to 52% during the period August to November 2021 when the Delta variant was dominant (>99% in this period), and from 46 to 67% during weeks 48 2021 to week 1 2022 when both variants were circulating. Variant detection dropped to 18% in week 2 of 2022 when Omicron became the dominant variant accounting for more than 96% of the screened cases (Figure 1). During the first weeks when Omicron was first detected, most Omicron cases were reported from the counties of Oslo and Viken. Overall, 17,103 infections were identified as Delta and 5,121 as Omicron (including 135 sub-variant BA.2 cases that were reported in weeks 1-2 of 2022). Further details on characteristics of the study population are presented in Table 1.

**Table 1:** Characteristics of the study population and of those who tested positive for SARS-CoV-2 Delta and Omicron variant, Norway, 25 August 2021-16 January 2022 (n = 372,179 individuals).

| Characteristics | Study population<sup>a</sup><br>(n = 372,179) |  | Reported infections |  |  |  |  |  |
| --- | --- | --- | --- | --- | --- | --- | --- | --- |
|  |  |  | All infections<br>(n=52,891) |  | Delta variant<br>(n=17,103) |  | Omicron variant<br>(n=5,121) |  |
|  | n | % | n | % | n | % | n | % |
| **Sex** |  |  |  |  |  |  |  |  |
| Male | 191,316 | 51.4 | 26,967 | 51.0 | 8,681 | 50.8 | 2,573 | 50.2 |
| Female | 180,863 | 48.6 | 25,924 | 49.0 | 8,422 | 49.2 | 2,548 | 49.8 |
| **Age groups in years** |  |  |  |  |  |  |  |  |
| 12-15 | 254,498 | 68.4 | 37,993 | 71.8 | 11,738 | 68.6 | 4,065 | 79.4 |
| 16-17 | 117,681 | 31.6 | 14,898 | 28.2 | 5,365 | 31.4 | 1,056 | 20.6 |
| **Country of birth** |  |  |  |  |  |  |  |  |
| Norway | 329,590 | 88.6 | 45,149 | 85.4 | 14,623 | 85.5 | 4,337 | 84.7 |
| Outside of Norway | 42,583 | 11.4 | 7,742 | 14.6 | 2,480 | 14.5 | 784 | 15.3 |
| Unknown | 6 | 0.0 | 0 | 0.0 | 0 | 0.0 | 0 | 0.0 |
| **Risk of severe COVID-19<sup>b</sup>** |  |  |  |  |  |  |  |  |
| Low | 342,775 | 92.1 | 48,987 | 92.6 | 15,853 | 92.7 | 4,737 | 92.5 |
| Medium | 27,308 | 7.3 | 3,691 | 7.0 | 1,177 | 6.9 | 362 | 7.1 |
| High | 2,096 | 0.6 | 213 | 0.4 | 73 | 0.4 | 22 | 0.4 |

<sup>a</sup> Note that the study population in the main analysis includes the individuals at the beginning of the study period (25 August 2021) and that the population in the sub analyses for the period November 2021 to January 2022 is different since prior infections are excluded and few people died.
<sup>b</sup> Risk of severe disease based on underlying comorbidities that are associated with a moderate or high risk of serious illness regardless of age. Details on the definitions used are provided in the Supplement, part 1.

**Figure 1:**
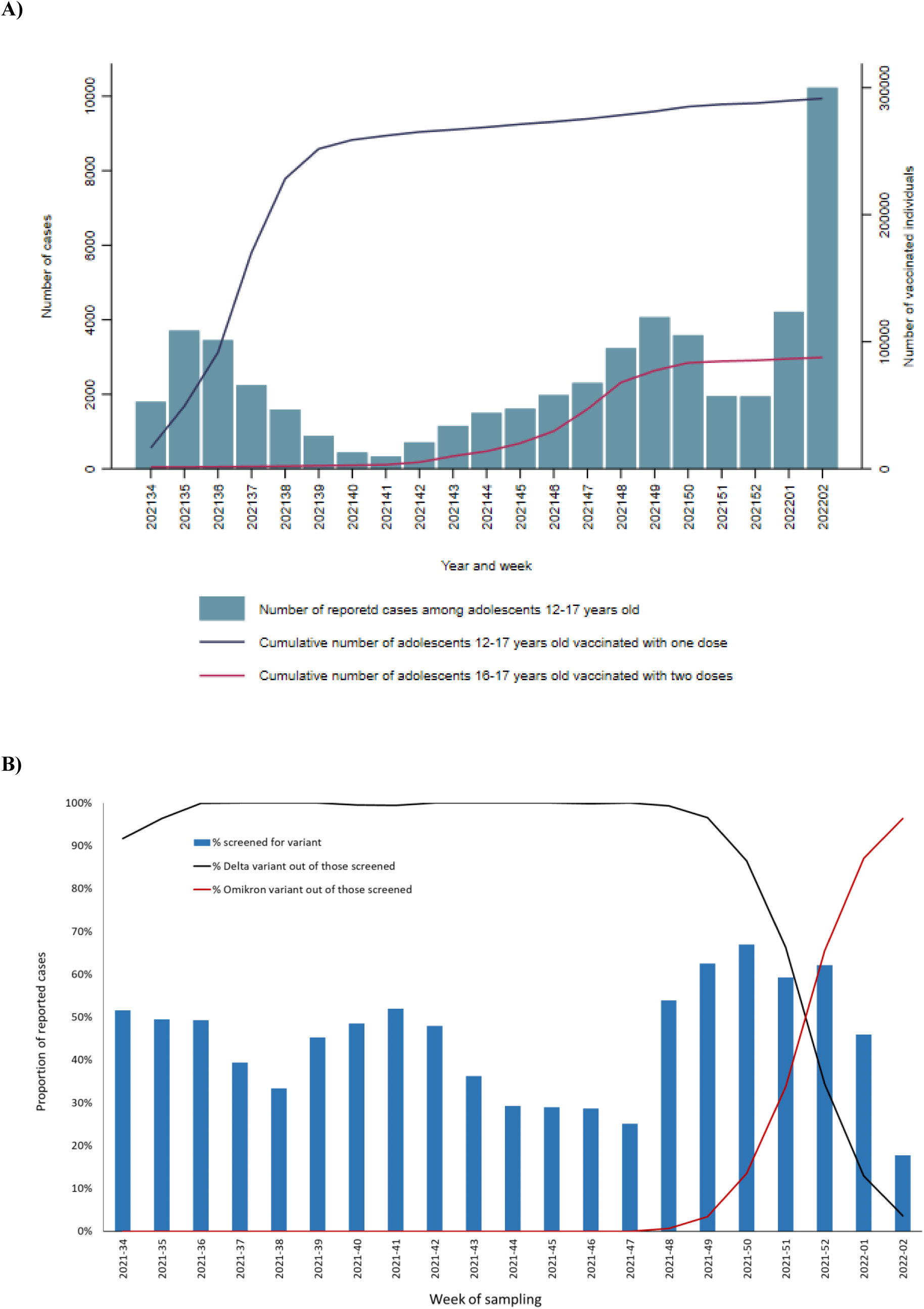
Number of COVID-19 reported cases by week of sampling and cumulative number of vaccinated adolescents 12-17 years old by vaccine doses received and week (panel A); proportion of reported COVID-19 cases among adolescents 12-17 years old with data on virus variant, and proportion of Delta and Omicron by week of sampling (panel B), Norway, 25 August 2021–16 January 2022 (n =52,891 cases).

Regarding vaccination coverage, just before the start of the study period (by the end of week 33), 3,445 adolescents 12-17 years old had received one dose of the vaccine and 1,121 adolescents 16-17 years old had received two doses. Vaccination uptake increased in the coming weeks and by week 48, when Omicron started circulating, 278,422 12-17-year-olds had received one dose of the vaccine and 67,955 16-17-year-olds two doses resulting in a vaccine coverage approximately 75% (278,422 /372,179) and 59% (67,955 /117,681) respectively for the relevant groups. By the end the study period, vaccine coverage with one dose among 12-17-year-olds was 78% (291,511/372,179) and with two doses among 16-17-year-olds was 74% (87,613/117,681) (Figure 1).

Testing activity seems to have been similar among unvaccinated adolescents and adolescents vaccinated with one or two doses throughout the study period, especially after week 38. The proportion of individuals tested per week ranged from 1·1 to 16·5% for the unvaccinated adolescents, 1·1 to 10·9% for those vaccinated with one dose and 1·0 to 8·7% for those vaccinated with two doses (Figure 2). Testing activity was similar for the two age groups, 12-15 and 16-17 years.

**Figure 2:**
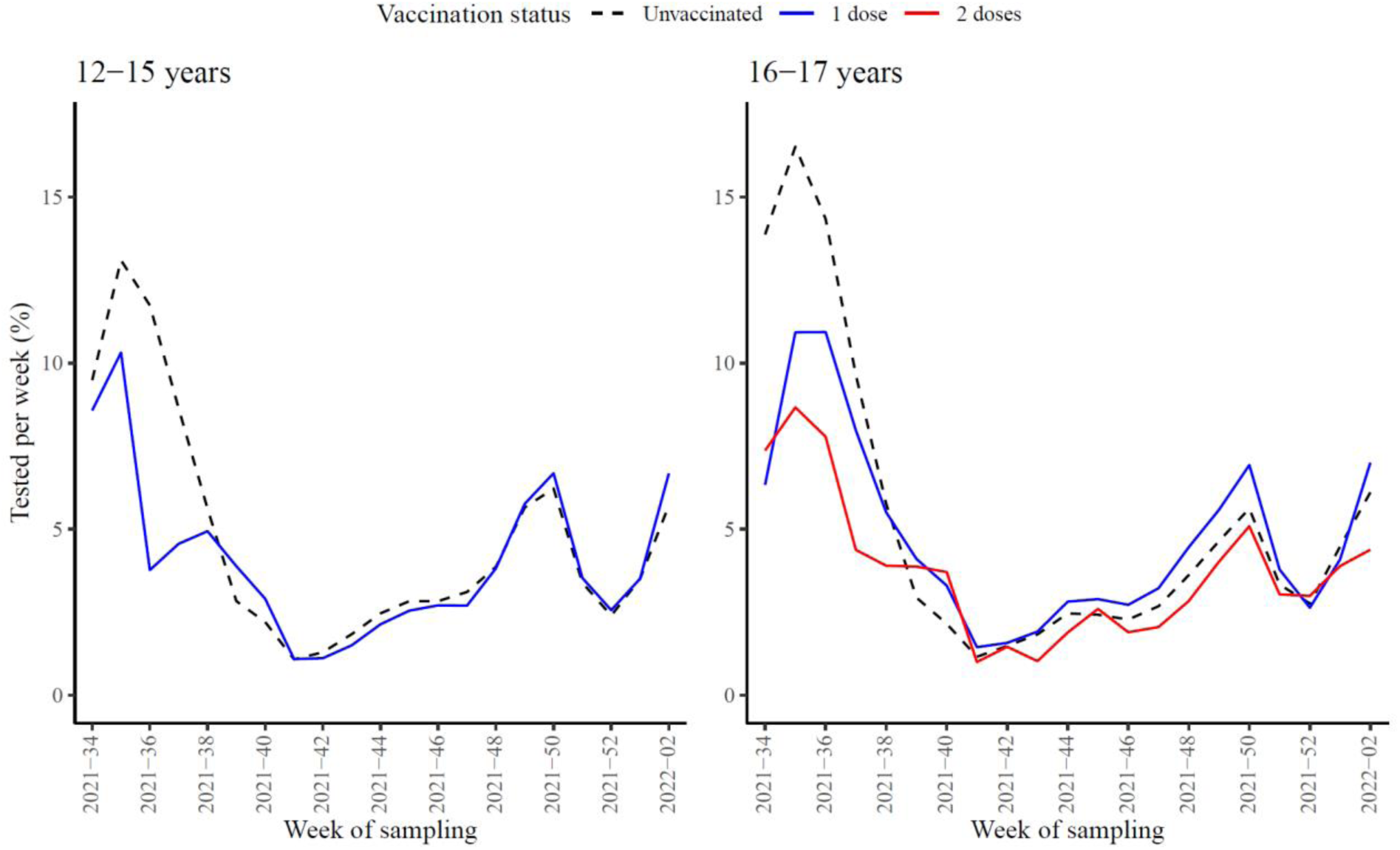
Proportion of individuals tested per week for the age group 12-15 (left panel) and 16-17 (right panel) years by vaccine status (unvaccinated: black dashed line, 1 dose: blue line, 2 doses: red line), Norway, 25 August 2021–16 January 2022 (n = 372,179 individuals).

### Vaccine effectiveness

#### Delta variant

Vaccine effectiveness against Delta infections was 68% (95% confidence interval [CI], 64–71%) and 62% (95%CI: 57–66%) in days 21-48 after the first vaccine dose among 12–15-year-olds and 16–17-year-olds respectively. After 77 days or more after the first dose, the vaccine effectiveness decreased to 49% (95%CI:46‒ 52%) and 48% (95%CI: 43–52%) respectively (figure 3, tableS2 in supplement). Among 16–17-year-olds that received two doses, the vaccine effectiveness was 91% (95%CI: 87‒93%) in days 7-34, 93% (95%CI: 90–95%) in days 35-62 and 84% (95%CI: 76–89%) in 63 days or more after the second dose.

**Figure 3:**
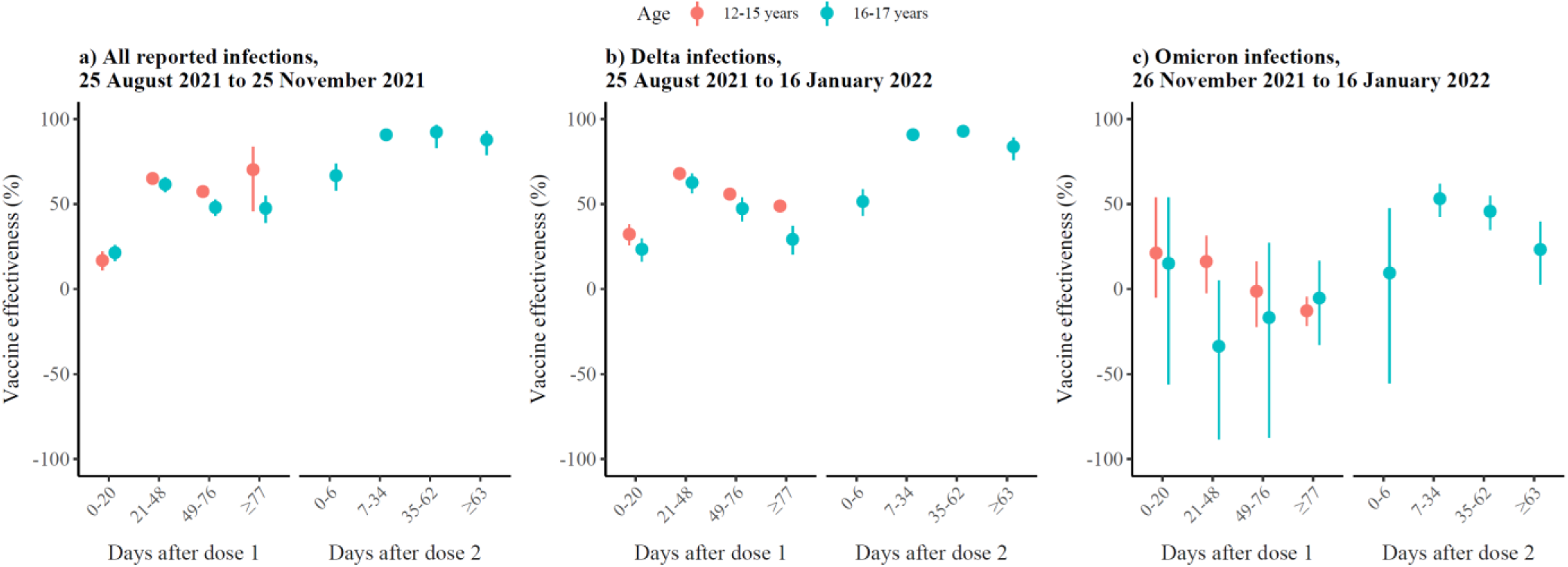
Adjusted vaccine effectiveness (aVE) for age 12-15 years (orange) and 16-17 (blue) by vaccination status against a) all reported infections from 25 August to 25 November, b) reported Delta infections from 25 August 2021 to 16 January 2022, and c) reported Omicron infections from 26 November to 16 January. We estimated aVE using Cox regression stratified by age, sex, underlying, county of residence, country of birth, crowding and underlying comorbidities. Note 1: The Vaccine effectiveness in 0-6 days after the second dose probably reflects the effect of the first vaccine dose. Note 2: Overall, during 26 November 2021 to 16 January 2022: There were 8,013 Delta infections reported, of which 8,011 were included in the Cox regression (two cases had date of event before date of entry due to errors). From those 8,011 Delta cases, the 6,507 were in age group 12-15 and 1,504 in the age group 16-17 years. Moreover, there were 5,123 Omicron infections of which 5,120 were included in the Cox regression (three cases had date of event before date of entry due to errors). From those 5,120 Omicron cases, the 4,064 were in age group 12-15 and 1,056 in the age group 16-17. Note 3: Details on the numbers presented here can be found in supplement, tableS2.

Our vaccine effectiveness estimates were similar to the above when we conducted our analysis using all reported infections (assuming that all are Delta) during the period 25 August 2021 to 25 November 2021 with slightly higher estimates 63 days or more after the second dose (88% vs 84% with overlapping CI). Also, the estimates for vaccine effectiveness 77 days or more after one dose were significantly higher when taking into account all infections than just those screened as Delta (48% vs 29% with no overlapping CI). This was possibly due to a longer follow-up period in the analysis using Delta cases (up to 16 January 2022) than the analysis for all infections (up to 25 of November 2021), and time since vaccination in the analysis of vaccine effectiveness 77 days or more after one dose was longer.

#### Omicron variant

For both age groups, we found no protection against Omicron SARS-CoV-2 infection after receiving one dose (figure 3, tableS2 in supplement). More specifically, no significant effect was found for all timepoints apart from the period of 77 days or more after the dose for which there was a negative vaccine effectiveness estimate for the 12–15-year-olds (−13%, 95% CI: -5%– -22%). Among 16–17-year-olds who received two doses, the vaccine effectiveness against Omicron infection peaked 7-34 days after the second dose at 53% (95%CI: 43–62%) and gradually decreased to 23% (95%CI: 3–40%) 63 days or more after the second dose. Overall, the protective effect of the vaccine against Omicron infection (when observed) was significantly lower than Delta in all timepoints apart from the period 0-6 days after two doses in 16-17-year-olds where the CI were slightly overlapping.

In our study, we estimated vaccine effectiveness against infection, and we did not include any vaccine effectiveness analyses against hospitalisation or death (severe disease) due to small numbers of events. During the study period, there were 31 hospitalisations with COVID-19 as main reason for admission, and 4 ICU admissions from which none died.

## Discussion

We found that two doses of BNT162b2 vaccine were highly effective against any (symptomatic or not) SARS-CoV-2 Delta infection and moderately effective against any Omicron infection among adolescents aged 16-17 years. One dose was moderately effective against Delta infection among adolescents aged 12-17 years, but we found no protection against Omicron infection. For the Omicron variant, waning immunity was observed faster than for Delta.

Vaccine effectiveness above 90% against the Delta variant 7-62 days after the second dose is in line with results from other studies among adolescents in England, Israel, and USA even though different study designs were used (6,9,10). A reduced effectiveness of the mRNA vaccines against infection with the Omicron variant compared to the Delta variant has been documented in several preprint studies, primarily in adults (14-16), but also in one preprint study in adolescents in England (9). In our study we found no protective effect after one dose, but the study from England among adolescents estimated that vaccine effectiveness against symptomatic Omicron infection after one dose peaked at 50% in days 14-20 after vaccination among 12-15-year-olds and at 53% in days 21-27 after vaccination among 16-17-year-olds. We found that the protective effect against any infection caused by Omicron peaked between 7-34 days and was 53% after the second dose for 16-17-year-olds, whereas the relevant highest vaccine effectiveness was 76% between 7-13 days after the second dose in the study from England. We should note here that evidence suggests higher rates of asymptomatic carriage for Omicron than other variants of concern (17), and vaccine effectiveness that includes asymptomatic cases is expected to be lower than when including symptomatic cases only. As the population, study design, interval between doses, the exact interval used for the assessment of vaccine effectiveness, outcomes (symptomatic or asymptomatic infections) and epidemic are site dependent, it is important to have data from various settings for guidance of vaccine policies.

Like in adults, we noticed that vaccine-induced protection against infection wanes with time for both variants (14, 18). Waning of protection against Delta infection among adolescents has been reported from other studies as well (6, 19). We found that protection against Omicron infection wanes faster compared to Delta after the second dose as it has also been reported in the preprint study from England (9). Prevention of infection likely requires maintaining high antibody titres, which could be achieved by booster vaccination. However, the protection against infection seems to be temporary and we have currently low certainty evidence available about the effectiveness of booster doses in adolescents 12-17 years(20, 21). As protection against severe disease is shown to wane less than protection against infection, the necessity to prevent infection in previously primary vaccinated adolescents is still not clear. In our study, we did not include any vaccine effectiveness analyses against hospitalisation or death (severe disease) due to small numbers of events. However, studies in adults have consistently shown that the VE against severe disease is higher than for infection and duration of protection is longer (14, 18). Therefore, it is reasonable to expect that this would also be the case for younger individuals.

A strength of this study is access and ability to link data collected from different national registries that allows us to estimate vaccine effectiveness at a national level for adolescents. The study also has some limitations. For the combined registries there can be errors for specific individuals, missing individuals, and older data (e.g county of residence, household crowding). Data used from these registries are not collected for the purpose of this study alone and some details might not be as precise. But there is no reason to believe that such discrepancies, if they exist, would differ among the different groups that we have compared. Various aspects related to disease dynamics, risk of infection, testing strategy, and response changed over the study period, making it challenging to identify all elements that affect vaccine effectiveness and risk of infection. However, we expect these changes to be similar between the adolescents in this study and by using time explicitly in our models we take some of these changes indirectly into consideration. However, residual confounding may still be present. The sensitivity analyses using data from only cases screened for variants compared to all reported cases provided similar estimates on vaccine effectiveness (supplementary material, part 2). Therefore, we conclude that using all reported SARS-CoV-2 cases in periods with one variant dominating can provide similar vaccine effectiveness estimates with larger power when only a small proportion of samples are screened.

Even though testing capacity is high in Norway and regular testing was recommended for adolescents in areas and periods with high infection rates that attended school during the study period, it is likely that not all individuals infected with SARS-CoV-2 have been detected. This could affect the estimates since the fraction of unidentified cases might differ by vaccine status. Moreover, the estimated vaccine effectiveness can be affected if the number and types of social contacts by an individual depends on his/her vaccination status (22), for example if getting vaccinated results in behavioural changes associated with higher risk of exposure (underestimate) and/or lower likelihood of testing (overestimate). We are not able to model these effects in our analyses. However, we expect in this study population that risk behaviour, type of contacts and testing would be relatively homogenous since this population had similar exposures (i.e. being at school), testing requirements did not depend on vaccination status and we did not observe any big differences in the proportion of individuals tested per week in the different vaccination status groups. We should also note that the vaccine effectiveness estimates against Omicron infection here are mainly against sub-variant BA.1 since they were estimated in a period when BA.1 was the predominating sub-variant.

In conclusion, one and two doses of BNT162b2 among adolescents protected well against Delta SARS-CoV-2 infection with higher protection among those with two doses, but the effectiveness waned with time since last dose. We found no protection against Omicron SARS-CoV-2 infection after one vaccine dose, and moderate effectiveness after two doses. For the Omicron variant, waning immunity was observed faster than for Delta. These results suggest that vaccination might have limited effect in the spread of the Omicron variant among adolescents, which should be considered when providing guidance on future vaccine policies in this group. In addition, the vaccine effectiveness should be re-evaluated when other variants appear as they might have different outcomes as shown between Delta and Omicron infections.

## Data Availability

The dataset analysed in the study contains individual‒level linked data from various central health registries, national clinical registries, and other national administrative registries in Norway. The researchers had access to the data through the national emergency preparedness registry for COVID‒19 (Beredt C19), housed at the Norwegian Institute of Public Health (NIPH). In Beredt C19, only fully anonymised data (i.e. data that are neither directly nor potentially indirectly identifiable) are permitted to be shared publicly. Legal restrictions therefore prevent the researchers from publicly sharing the dataset used in the NH study that would enable others to replicate the study findings. However, external researchers are freely able to request access to linked data from the same registries from outside the structure of Beredt C19, as per normal procedure for conducting health research on registry data in Norway. Further information on Beredt C19, including contact information for the Beredt C19 project manager, and information on access to data from each individual data source, is available at https://www.fhi.no/en/id/infectious-diseases/coronavirus/emergency-preparedness-register-for-covid-19/.

## Authors’ contributions

LV, JDB, SVW, MGI, and HM were involved in the conceptualisation of the study. LV, and HM drafted the study protocol with feedback from JDB, SVW, JS, MGI. LV, and HM coordinated the study. LV, JS, PL, and HM had directly assessed and verified the underlying data and produced figures and tables. HB cleaned the variant data. LV, JS, PL, HB, HM had access to the data and the rest of the authors had the opportunity to request access to the data for this study. LV conducted the statistical analysis in consultation with HM. All co-authors contributed to the interpretation of the results. LV drafted the manuscript with support from JDB, and HM. All co-authors contributed to the revision of the manuscript and approved the final version for submission. The corresponding author attests that all listed authors meet authorship criteria and that no others meeting the criteria have been omitted.

## Conflict of interest

The authors declare that they have no competing interests.

## Acknowledgements

We wish to thank all those who have helped to collect and report data to the national emergency preparedness registry at the Norwegian Institute of Public Health (NIPH) throughout the pandemic. We are grateful to all health professionals that contributed to vaccinating the Norwegian population and those performing millions of laboratory tests for COVID-19. Thanks also to the staff at the regional laboratories and the Virology and Bacteriology departments at NIPH involved in the analyses of samples, national variant identification and whole genome analysis of SARS-CoV-2 viruses. We would also like to acknowledge our colleagues at the NIPH who have contributed to the data cleaning from different registries throughout the pandemic.

## Notes

### Competing Interest Statement

The authors have declared no competing interest.

