## Supplementary material for "Vaccine effectiveness with BNT162b2 (Comirnaty, Pfizer-BioNTech) vaccine against reported SARS-CoV-2 Delta and Omicron infection among adolescents, Norway, August 2021 to January 2022"

### Supplementary appendix

### 1. Data sources and linkages

All data in this study came from the national emergency preparedness register, Beredt C19 [1]. Beredt C19 contains individual-level data from central health registries, national clinical registries and other national administrative registries. The data sources and variables used are shown in Table S1

**Table S1. Data sources in the Norwegian preparedness registry (BeredtC19) used in this study and variables retrieved from each source**

| Norwegian Abbreviation | Full name of data source | Information obtained |
|---|---|---|
| **DSF** | The National Population Register | Age, Sex, County of residence*, Country of birth, Date of death |
| **SYSVAK** | The National Immunisation Register | Date of vaccination, Vaccine product type |
| **NIPaR** | Norwegian Intensive Care and Pandemic Registry | Date of hospitalisation, COVID-19 as main cause of admission, Date of ICU admission |
| **MSIS** | The Surveillance System for Infectious Diseases | Date of sample of SARS-CoV-2 positive test, Date of COVID-19 associated death |
| **SSB** | Statistics Norway | "living crowded" (see definition below) |
| **Beredt C19 risikogrupper** | Table prepared in BeredtC19 Source: Norwegian Patient Registry (NPR): individual level data from all public specialist health-care services in Norway. | Defines risk groups (see definition below) |

** The county of residence was updated in January 2022, which might lead to some errors for individuals who have moved the last half year*

Reported cases of COVID-19, reinfections and deaths: We included data on reported cases of laboratory-confirmed SARS-CoV-2 infection and deaths from the Norwegian Surveillance System for Communicable Diseases (MSIS). As of January 2022, in MSIS, reinfections are registered if there are ≥6 months between two positive sampling dates for an individual, although this will exclude reinfections within a 6-month period, of which the Omicron variant could be of higher risk.

Laboratory testing for variants: Data on virus variants came from the MSIS laboratory database (national laboratory database), which receives SARS-CoV-2 test results from all Norwegian microbiology laboratories [2].

**Additional definitions used:**

<u>Crowding</u>: Individuals are considered to live in crowded conditions if the number of rooms is lower than the number of residents or one resident lives in one room, and the number of square metres (P-area) is below 25 sq. m. per person. If the number of rooms or the P-area is not specified, a household will be regarded as crowded if one of these criteria is met [3].

<u>Risk groups:</u> Some underlying medical conditions increase the risk of severe COVID-19 outcomes, regardless of age. These individuals have been prioritised in the vaccination campaigns in Norway. The underlying comorbidities that have been defined as increasing the risk of severe COVID-19 are divided into two groups:

High risk: people with diseases/conditions that carry a high risk of severe COVID-19:

- Organ transplant
- Immunodeficiency
- Haematological cancer in the last five years
- Other active cancers
- Neurological or neuromuscular diseases that cause impaired cough or lung function (e.g., ALS and cerebral palsy)
- Chronic kidney disease, or significant renal impairment.

Medium risk: people with diseases/conditions that entail a moderate risk of severe COVID-19:

- Chronic liver disease or significant hepatic impairment
- Diseases requiring immunosuppressive therapy
- Diabetes
- Chronic lung disease including cystic fibrosis and severe asthma which have required the use of high dose inhaled or oral steroids within the past year
- Obesity with a body mass index (BMI) of ≥35 kg/m2
- Dementia
- Chronic heart and vascular disease (with the exception of high blood pressure) and stroke

### 2. Vaccine effectiveness

#### 2.1 Statistical analyses, including sensitivity analyses

We estimated the vaccine effectiveness against all infections (Delta- and Omicron infections in separate analyses), using Cox proportional hazards models with vaccination status as a time-dependent covariate, and with explicit time to account for changes in the baseline hazard over time. Vaccine status is implemented as a regular parameter and assumes proportionality with vaccine effectiveness defined as 100*(1 $-\beta$), with $\beta$ the proportional hazard associated with vaccine status. Using stratification to adjust for the covariates allow each combination of covariate groups to have their own baseline hazard rate, while still assuming that vaccination leads to a proportional change in risk of infection. In all our adjusted vaccine effectiveness estimates presented here, we stratified for sex, country of birth, county of residence, crowding and underlying comorbidities associated with increased risk of severe COVID-19.

More specifically, we estimated the vaccine effectiveness among individuals with no prior SARS-CoV-2 infection after receiving one dose for individuals 12-15 years old and after one or two doses for individuals 16-17 years old against a) all reported infections from 25 August to 25 November, b) reported Delta infections from 25 August 2021 to 16 January 2022 and c) reported Omicron infections from 26 November to 16 January. The analyses for all reported cases during the period 25 August to 25 November is conducted assuming that all reported cases were Delta since Delta accounted for nearly 100 % of all samples analysed for genetic variant in that period. We restricted the vaccine effectiveness analyses for Omicron infection after the first case of Omicron was reported in adults on the 26 of November 2021. We closed the follow up period on 16 January since the comprehensive screening activity ceased as Omicron reached more than 90% prevalence.

In the Cox proportional hazards models used, unvaccinated person-time included days before receiving the first dose of a COVID-19 vaccine (if received) and vaccinated person-time included number of days following the receipt of one or two doses of the vaccine (based on the vaccination programme). For adolescents that got infected, the end/event time for these analyses was the first sampling date where the participants tested positive (right censored). We also right-censored individuals at the time of death (all cause), date of third dose or end of follow-up period (25 of November or 16 of January according to the last day for the three outcomes).

We present here the results from our adjusted vaccine effectiveness analyses in the tables S2, below. In addition, we present the results from the sensitivity analyses that were performed in table S3 to evaluate the impact in estimating vaccine effectiveness for a specific variant when only including (using data) on screened cases since usually only part of all reported cases is screened for variant.

Table S2: Adjusted vaccine effectiveness (aVE) by age group and vaccination status against a) all reported infections from 25 August to 25 November, b) reported Delta infections from 25 August 2021 to 16 January 2022 and c) reported Omicron infections from 26 November to 16 January. We estimated aVE using Cox regression stratified by age, sex, underlying, county of residence, country of birth, crowding and underlying comorbidities.

**Table S2: Adjusted vaccine effectiveness (aVE) by age group and vaccination status against a) all reported infections from 25 August to 25 November, b) reported Delta infections from 25 August 2021 to 16 January 2022 and c) reported Omicron infections from 26 November to 16 January. We estimated aVE using Cox regression stratified by age, sex, underlying, county of residence, country of birth, crowding and underlying comorbidities.**

| | **Age group 12-15** | | **Age group 16-17** | |
|---|---|---|---|---|
| | **Number of cases** | **Adjusted* vaccine effectiveness (95% CI)** | **Number of cases** | **Adjusted* vaccine effectiveness (95% CI)** |
| **All infections, 25 August to 25 November 2021** | | | | |
| Unvaccinated | 9,800 | Ref | 4,739 | Ref |
| 1st dose, 0-20 days | 1,506 | 16·8 (11·12–22·0) | 2,075 | 21·4 (16·4–26·0) |
| 1st dose, 21-48 days | 952 | 65·0 (62·3–67·6) | 517 | 61·5 (57·1–65·5) |
| 1st dose, 49-76 days | 1,601 | 57·3 (54·4–60·0) | 1,305 | 48·0 (43·3–52·4) |
| 1st dose, ≥77 days | 11 | 70·2 (45·9–83·6) | 279 | 47·5 (39·0–54·9) |
| 2nd dose, 0-6 days | NA | NA | 78 | 66·7 (57·9–73·7 |
| 2nd dose, 7-34 days | NA | NA | 45 | 90·7 (87·4–93·1) |
| 2nd dose, 35-62 days | NA | NA | 6 | 92·3 (82·9–96·6) |
| 2nd dose, ≥63 days | NA | NA | 13 | 87·8 (78·8–92·9) |
| **Delta infections, 25 August 2021 to 16 January 2022** | | | | |
| Unvaccinated | 6,783 | Ref | 2,771 | Ref |
| 1st dose, 0-20 days | 665 | 32·3 (25·9–38·2) | 929 | 23·4 (16·3–29·8) |
| 1st dose, 21-48 days | 379 | 67·9 (64·0–71·4) | 246 | 62·6 (56·2–68·0) |
| 1st dose, 49-76 days | 1,257 | 55·8 (52·7–58·8) | 461 | 47·3 (40·0–53·8) |
| 1st dose, ≥77 days | 2,640 | 48·8 (46·0–51·5) | 515 | 29·3 (20·4–37·1) |
| 2nd dose, 0-6 days | NA | NA | 216 | 51·4 (43·2–58·5) |
| 2nd dose, 7-34 days | NA | NA | 164 | 90·8 (89·1–92·3) |
| 2nd dose, 35-62 days | NA | NA | 34 | 92·8 (89·8–94·9) |
| 2nd dose, ≥63 days | NA | NA | 29 | 83·7 (75·9–89·0) |
| **Omicron infections, 26 November 2021 to 16 January 2022** | | | | |
| Unvaccinated | 973 | Ref | 214 | Ref |
| 1st dose, 0-20 days | 49 | 21·2 (-5·1–53·8) | 11 | 15·1 (-56–53·8) |
| 1st dose, 21-48 days | 108 | 16·2 (-2·4–31·3) | 39 | -33·7 (-88·3–5·1) |
| 1st dose, 49-76 days | 125 | -1·3 (-22·4–16·2) | 19 | -16·8 (-87·3–27·1) |
| 1st dose, ≥77 days | 2,779 | -12·8 (-21·7– -4·6) | 110 | -5·3 (-32·9–16·6) |
| 2nd dose, 0-6 days | NA | NA | 15 | 9·5 (-55·2–47·3) |
| 2nd dose, 7-34 days | NA | NA | 204 | 53·1 (42·6–61·7) |
| 2nd dose, 35-62 days | NA | NA | 330 | 45·7 (34·8–54·7) |
| 2nd dose, ≥63 days | NA | NA | 114 | 23·3 (2·7–39·5) |

Note 1: The Vaccine effectiveness in 0-6 days after the second dose probably reflects the effect of the first vaccine dose.
Note 2: Overall, during 26 November 2021 to 16 January 2022: There were 8,013 Delta infections reported, of which 8011 were included in the Cox regression (two cases had date of event before date of entry due to errors). From those 8,011 Delta cases, the 6,507 were in age group 12-15 and 1,504 in the age group 16-17 years. Moreover, there were 5,123 Omicron infections of which 5,120 were included in the Cox regression (three cases had date of event before date of entry due to errors). From those 5,120 Omicron cases, the 4,064 were in age group 12-15 and 1,056 in the age group 16-17.

**Table S3: Adjusted vaccine effectiveness (aVE) by age group and vaccination status against a) all reported infections from 25 August to 25 November and b) reported Delta infections from 25 August to 25 November2021. We estimated aVE using Cox regression stratified by age, sex, underlying, county of residence, country of birth, crowding and underlying comorbidities.**

| | Age group 12-15 | | Age group 16-17 | |
|---|---|---|---|---|
| | **Number of cases** | **Adjusted* vaccine effectiveness (95% CI)** | **Number of cases** | **Adjusted* vaccine effectiveness (95% CI)** |
| **All infections** | | | | |
| Unvaccinated | 9,800 | Ref | 4,739 | Ref |
| 1st dose, 0-20 days | 1,506 | 16·8 (11·12–22·0) | 2,075 | 21·4 (16·4–26·0) |
| 1st dose, 21-48 days | 952 | 65·0 (62·3–67·6) | 517 | 61·5 (57·1–65·5) |
| 1st dose, 49-76 days | 1,601 | 57·3 (54·4–60·0) | 1,305 | 48·0 (43·3–52·4) |
| 1st dose, ≥77 days | 11 | 70·2 (45·9–83·6) | 279 | 47·5 (39·0–54·9) |
| 2nd dose, 0-6 days | NA | NA | 78 | 66·7 (57·9–73·7 |
| 2nd dose, 7-34 days | NA | NA | 45 | 90·7 (87·4–93·1) |
| 2nd dose, 35-62 days | NA | NA | 6 | 92·3 (82·9–96·6) |
| 2nd dose, ≥63 days | NA | NA | 13 | 87·8 (78·8–92·9) |
| **Delta variant** | | | | |
| Unvaccinated | 3,945 | Ref | 2,233 | Ref |
| 1st dose, 0-20 days | 530 | 27·5 (19·1–35·0) | 889 | 23·6 (16·3–30·3) |
| 1st dose, 21-48 days | 285 | 68·9 (64·3–72·9) | 227 | 63·3 (56·5–69·0) |
| 1st dose, 49-76 days | 463 | 56·0 (50·3–61·0) | 398 | 49·4 (41·0–56·7) |
| 1st dose, ≥77 days | 3 | 70·1 (6·2–90·5) | 68 | 46·5 (27·8–60·3) |
| 2nd dose, 0-6 days | NA | NA | 24 | 62·5 (42·9–75·4) |
| 2nd dose, 7-34 days | NA | NA | 14 | 89·9 (82·8–94·1) |
| 2nd dose, 35-62 days | NA | NA | 0 | – |
| 2nd dose, ≥63 days | NA | NA | 8 | 80·3 (60·0–90·3) |

Note: Overall, during 25 August 2021 to 25 November 2021: There were 22,937 all infections of which 22,935 were included in the Cox regression (two cases had date of event before date of entry due to errors). From those 22,935 all infections, the 13,878 were in age group 12-15 and 9,057 in the age group 16-17. There were 9,092 Delta infections reported, of which 9,090 were included in the Cox regression (two cases had date of event before date of entry due to errors). From those 9,090 Delta cases, the 5,229 were in age group 12-15 and 3,861 in the age group 16-17 years.
